# Brain Injury after Transcatheter Aortic Valve Replacement for Bicuspid Versus Tricuspid Aortic Valve Stenosis

**DOI:** 10.1101/2020.02.19.20023184

**Authors:** Jiaqi Fan, Xian Fang, Chunhui Liu, Gangjie Zhu, Cody R. Hou, Jubo Jiang, Xinping Lin, Lihan Wang, Yuxin He, Qifeng Zhu, Stella Ng, Zexin Chen, Haitao Hu, Xianbao Liu, Jian’an Wang, Martin B. Leon

**Affiliations:** Department of Cardiology, Second Affiliated Hospital Zhejiang University School of Medicine, Hangzhou, People’s Republic of China; Zhejiang University School of Medicine, Hangzhou, People’s Republic of China; Department of Cardiology, Lanxi People’s Hospital, Lanxi, People’s Republic of China; College of Biological Sciences, University of Minnesota Twin Cities, Minneapolis, MN, United States of America; Department of Clinical Epidemiology and Biostatistics, Second Affiliated Hospital Zhejiang University School of Medicine, Hangzhou, People’s Republic of China; Department of Neurology, Second Affiliated Hospital Zhejiang University School of Medicine, Hangzhou, People’s Republic of China; Division of Cardiology, Columbia University Medical Center and the Cardiovascular Research Foundation, New York, New York

**Keywords:** Brain Injury, Cerebral Ischemic Lesions, Transcatheter Aortic Valve Replacement, Bicuspid Aortic Valve

## Abstract

**OBJECTIVES:** This study sought to evaluate the risk of brain injury in bicuspid aortic valve (BAV) patients following transcatheter aortic valve replacement (TAVR).

**BACKGROUND:** An increasing number of BAV patients are undergoing TAVR, but the risk of brain injury in diffusion-weighted magnetic resonance imaging (DW-MRI) is currently unknown.

**METHODS:** A total of 204 consecutive severe aortic stenosis patients who underwent TAVR were enrolled. 83 (40.7%) patients were BAV patients and the other 121 patients were tricuspid aortic valve (TAV) patients. All patients received DW-MRI at baseline, 2 to 7 days after TAVR.

**RESULTS:** Mean ages (mean ± SD: 75.8 ± 6.7 years vs. 78.9 ± 6.6 years, p = 0.004) and STS scores (6.0 ± 3.7 vs. 7.1 ± 4.2, p = 0.044) of the BAV and TAV patients were significantly different, while the overt stroke rates (2.4% vs. 1.7%, p = 0.704) were comparable between two groups. BAV patients were associated with higher number of new lesions (5.69 ± 6.22 vs. 3.50 ± 4.16, p = 0.008), total lesion volume [median(interquartile range): 290(70-930) mm^3^ vs. 140(35-480) mm^3^, p = 0.008], and the volume per lesion [70.0(45.0-115.0) mm^3^ vs. 57.5(24.5-93.0) mm^3^, p = 0.037] in DW-MRI. Moreover, the proportion of patients with lesions larger than 1cm^3^ (28.6% vs. 10.9%, p = 0.005) and the number of new lesions in the middle cerebral arteries zone (1.46 ± 2.07 vs. 0.98 ± 1.84, p = 0.039) and intermediate zone between the anterior cerebral and middle cerebral arteries (ACA/MCA) (1.07 ± 1.68 vs. 0.50 ± 1.05, p = 0.007), and between the vertebral artery and basilar artery (VA/BA) (1.01 ± 1.35 vs. 0.77 ± 1.44, p = 0.033) were higher in BAV patients than in TAV patients.

**CONCLUSIONS:** BAV patients may encounter more severe brain injuries not only due to greater number of lesions but also due to larger lesion size, especially in the ACA/MCA, MCA and VA/BA lesions zone.

## Introduction

Transcatheter aortic valve replacement (TAVR) is an established therapy in symptomatic severe aortic stenosis (AS) patients at prohibitive, high and moderate risk for surgical aortic valve replacement (1–6). The US Food and Drug Administration (FDA) has expanded the indication to include severe AS patients who are at low risk for death or major complications associated with open-heart surgery for several transcatheter heart valves (7, 8). Although many randomized clinical trials for TAVR show excellent outcomes, bicuspid aortic valve (BAV) stenosis was excluded from these clinical trials due to its unique morphological features (1–9). Only a few small series and registry studies demonstrated the safety and efficacy of TAVR in BAV patients (10–14). Recently several studies have demonstrated a high incidence of new cerebral ischemic lesions on post-TAVR diffusion-weighted magnetic resonance imaging (DW-MRI) (15–17). However, there is no study has precisely assessed the number, volume, and distribution of the new cerebral ischemic lesions on post-TAVR DW-MRI in the BAV patients. Therefore, in the present study, we aim to compare brain injury after TAVR between BAV and TAV stenosis by post-TAVR DW-MRI.

## Methods

### Study Design and Patient Population

The TORCH (Transcatheter Aortic Valve Replacement Single Center Registry in Chinese Population, ClinicalTrials.gov Identifier: NCT02803294) registry is a single-center registry in Chinese population. The registry was initiated in June 2016, and BAV patients were also included in this registry. We collected data retrospectively from the TORCH registry. The study was approved by the medical ethics committee of Second Affiliated Hospital of Zhejiang University and carried out according to the principles of the Declaration of Helsinki. All patients provided written informed consent for TAVR and the use of anonymous clinical, procedural, and follow-up data for research.

For this study, we consecutively collected all severe AS patients greater than 18 years of age treated with transfemoral TAVR determined by an interdisciplinary heart team from December 1, 2016 to December 31, 2019 in the TORCH registry. Exclusion criteria: (a) implantation of incompatible metallic prosthesis or foreign body contraindicated to the DW-MRI examination, including pacemaker implantation; (b) history of a stroke or transient ischemic attack (TIA) within the prior six months; (c) absence of DW-MRI examination for other reasons, including in hospital death, conversion to surgical aortic valve replacement, and unplanned Cardiopulmonary Bypass before the DW-MRI examination, intolerance due to clinical situation, and refusal of the DW-MRI examination or overscheduling; and (d) poor quality of imaging or out of window period cannot be analyzed. All patients completed a 30-day follow-up.

Patients underwent a standard screening algorithm including echocardiography and multi-slice computed tomography (MSCT). Aortic annulus size was measured by multi-slice computed tomography in 3mensio software (3mensio Medical Imaging BV, the Netherlands). The threshold for detecting aortic root calcification was set at 650 HU; then, calcium volume was measured within the region from the left ventricular outflow tract (LVOT) to the leaflet tips. Aortic calcification was estimated visually and graded semi-quantitatively as follows: grade 0, no calcification; grade I, small non-protruding calcification less than 2 mm; Grade II, protruding calcifications more than 2 mm or calcification involving more than 50% of the valve circumference; Grade III, protruding calcifications more than 2 mm and involving more than 50% of the circumference; Grade IV, calcifications involving nearly 100% of the circumference (18). The transvalvular mean gradient, effective orifice valve area, and max transvalvular velocity were measured by transthoracic echocardiography (TTE) (18).

TAVR procedures were performed in our hybrid operating room as previously reported (19, 20). Unfractionated heparin was used in all procedures (50-70U/kg) to maintain an activated clotting time greater than 250 seconds. General anesthesia (GA) or local anesthesia with sedation was used during the procedure based on the evaluation from the anesthetist. Balloon valvuloplasty and post-dilatation were employed according to operator discretion. A large proportion of patients were implanted with self-expanding valves, such as: CoreValve (Medtronic Inc., Minnesota, USA), VenusA-Valve (Venus Medtech, Hangzhou, China), VitaFlow (Microport, Shanghai, China) and Taurus One-Valve (Peijia Medical, Suzhou, China). The rest of the patients were implanted with the Lotus valve (Boston Scientific, Marlborough, MA) or Edwards SAPIEN XT or SAPIEN 3 valve (Edwards Lifesciences, Irvine, California). Almost all patients were treated with dual antiplatelet therapy (aspirin 75 mg and clopidogrel 75 mg) with no indication of anticoagulation; when anticoagulation treatment was indicated, patients received warfarin or new oral anticoagulants.

### Bicuspid Aortic Valve

Bicuspid aortic valve morphology was classified as BAV or TAV according to the number of cusps and presence of raphes by Sievers and Schmidtke. Type 0 was assigned to morphologies characterized by the presence of 2 symmetric cusps and 1 commissure without evidence of a raphe. Type 1 was assigned to valve morphologies with 1 raphe, and type 2 when 2 raphes were present. Two authors reviewed and subsequently confirmed the diagnosis and classification of bicuspid AS in MSCT imaging before TAVR.

### Brain Magnetic Resonance Imaging

All patients received brain DW-MRI before and within 5.7 ± 2.8 days post TAVR procedure in the hospital using a 1.5-Tesla or 3-Tesla whole body MRI system (GE Signa). The imaging protocol was comprised of transversal T2-weighted turbo spin echo (TSE); 1.5-T: Repetition time (TR)/Echo time(TE): 4800/100ms; 3-T: TR/TE: 3300/80ms and transversal Fluid Attenuated Inversion Recovery (FLAIR); 1.5-T: TR/TE: 6000/120ms; 3-T: TR/TE: 1200/140ms. DWI was performed with a spin-echo echo-planar pulse sequence (1.5T: TE: 78ms; TR: 2921ms; matrix: 128×256; section thickness: 5mm; intersection gap: 1mm; total acquisition time: 21.4s; 3T: TE: 47ms; TR: 3866ms; matrix: 128×256; section thickness: 5mm; intersection gap: 1mm; total acquisition time: 46.3s;) with diffusion sensitization b-values of 0 and 1000s/mm^2^. Apparent diffusion coefficient (ADC) maps were calculated to identify findings with restricted diffusion. A new lesion was defined as a focal hyperintense area detected by the fluid-attenuated inversion recovery sequence, corresponding to a restricted diffusion signal in the diffusion-weighted imaging sequence, and confirmed by apparent diffusion coefficient mapping to rule out a shine-through artifact. The brain magnetic resonance imaging for new ischemic lesions was analyzed by the two independent authors with the software and confirmed by the neurological physician by MRIcron software. Vascular territories were classified according to previous studies (21, 22): anterior cerebral artery (ACA), middle cerebral artery (MCA), and posterior cerebral artery (PCA) for each side, respectively. Furthermore, vascular border zones (watershed zones) were defined as the area between ACA and MCA (ACA/MCA), MCA and PCA (MCA/PCA), vertebral artery and basilar artery (VA/BA)

### Data Collection

Data collection included baseline characteristics, procedural data, and pre-discharge outcomes. Baseline characteristics consisted of baseline clinical, laboratory, echocardiographic, and computed tomographic data, while pre-discharge outcomes were obtained from the local hospital database and assessed for quality. The incidence of new ischemic lesions, the number of lesions, total lesion volume, and total lesion volume/number of lesions before discharge in DW-MRI were compared between the two groups. Clinical outcomes were mortality, stroke (including disabling stroke and non-disabling stroke) and other clinical events before discharge according to the Valve Academic Research Consortium (VARC)-2 criteria(23). VARC-2 defines disabling stroke by a modified Rankin Scale (mRS) score of 2 or more at 90 days and an increase in at least one mRS category from an individual’s pre-stroke baseline. A VARC-2 non-disabling stroke is defined as an mRS score of <2 at 90 days or one that does not result in an increase in at least one mRS category from an individual’s pre-stroke baseline. The assessment of the mRS was used to maximize the detection of new or recurrent strokes, assist in the ongoing evaluation of events previously determined as TIAs, and provide an accepted and reliable indicator of the long-term impact of a given stroke. Multiple Pre-dilatation was defined as balloon pre-dilatation more than 1 time during the TAVR procedure. All data was stored in the database of TORCH registry and can be traced to the source.

### Statistical Analysis

Continuous variables following normal distribution were presented as mean ± SD and compared using the unpaired-sample Student’s *t*-test. Otherwise, skewed variables were compared using the Mann-Whitney U test. Categorical data were presented as count (percentages) and compared with the chi-square test. Some variables were presented as median (interquartile range, IQR) when there existed an outlier. Linear regression analysis was used to identify the relationship between patient and procedural factors and the number of new ischemic lesions. Univariate analysis was used to identify individual factors. Variables with a univariate significance of p < 0.1 and clinical factors from previous studies were entered into a multiple stepwise regression analysis to perform the multivariate linear regression analysis (24). A p < 0.05 was considered statistically significant. Statistical analysis was performed using SPSS software (version 20.0, SPSS Inc., Chicago, Illinois) and the figures were created in GraphPad Prism (version 6.0, GraphPad Software, San Diego, California).

## Results

A total of 204 patients were included in this study, with 83 BAV and 121 TAV patients. The BAV group consisted of 56 Type 0 patients and 27 Type 1 patients. TAV patients were older than BAV patients (75.8 ± 6.7 years vs. 78.8 ± 6.6 years, p = 0.004) and had higher STS scores (6.0 ± 3.6 vs. 7.1 ± 4.2, p = 0.044). TAV patients had a higher proportion of prior history of stroke (1.2% vs. 8.3%, p = 0.030). Patient selection flow was showed in **Figure 1**. All baseline demographics were summarized in **Table 1**.

**Figure 1.**
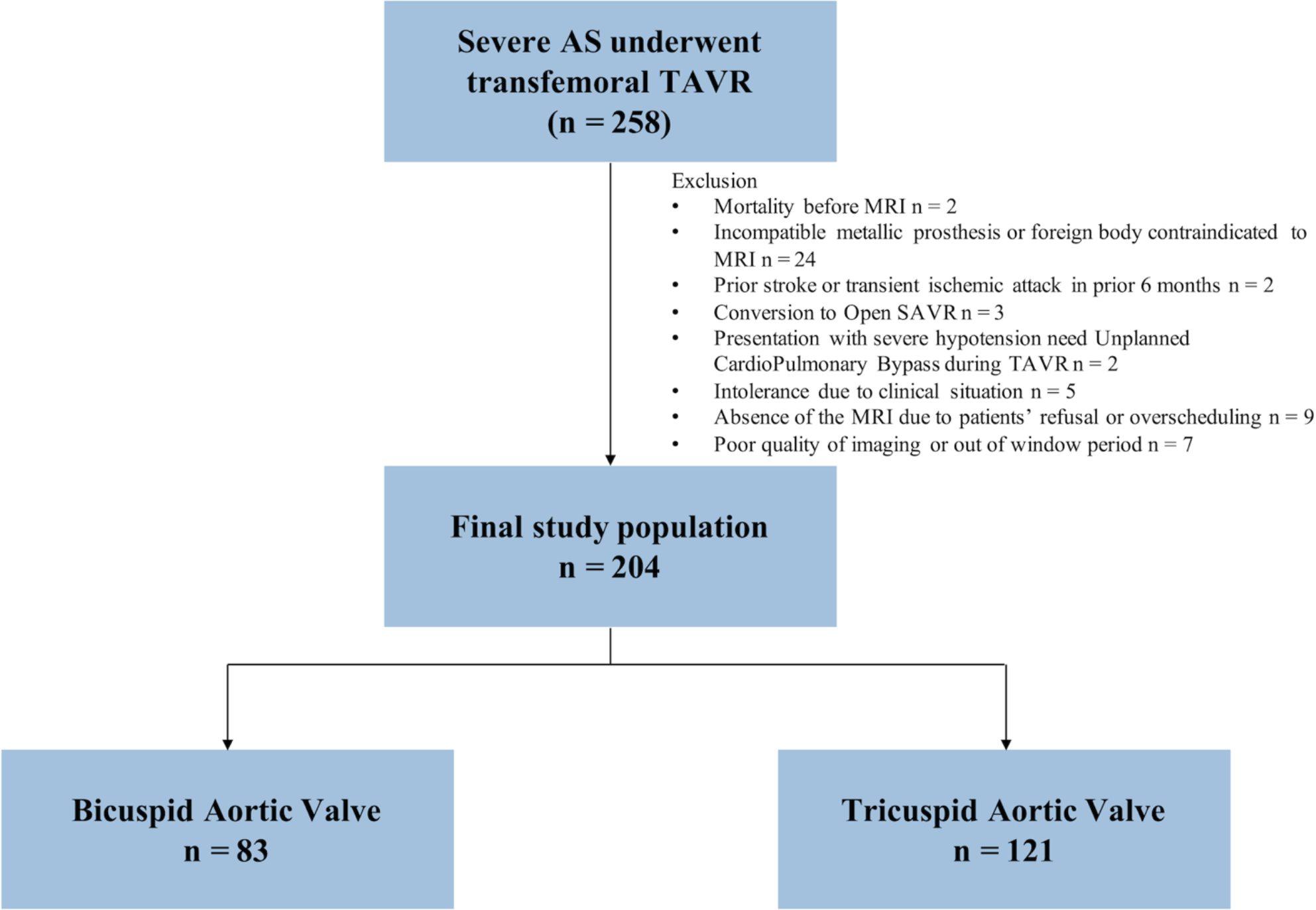
Patient selection flow.

**Table 1.** Baseline characteristics of BAV and TAV patients

|  | BAV |  |  | TAV | P value |
| --- | --- | --- | --- | --- | --- |
|  | BAV (Type 0)<br>n = 56 | BAV (Type 1)<br>n = 27 | Total<br>n = 83 | n = 121 |  |
| Age (yrs) | 75.1±7.0 | 77.3±6.1 | 75.8±6.7 | 78.8±6.6 | **0.004\*** |
| Male | 33(58.9) | 16(59.3) | 49(59.0) | 75(62.0) | 0.770 |
| BMI (kg/m<sup>2</sup>) | 22.28±2.91 | 21.96±2.58 | 22.17±2.79 | 23.02±3.56 | 0.058 |
| NYHA III/IV | 51(91.1) | 24(88.9) | 75(90.4) | 104(86.0) | 0.391 |
| STS | 5.74±3.64 | 6.54±3.65 | 6.00±3.64 | 7.11±4.21 | **0.044\*** |
| Smoker | 11(19.6) | 4(14.8) | 15(18.1) | 21(17.4) | 1.000 |
| Diabetes Mellitus | 14(25.0) | 10(37.0) | 24(28.9) | 29(24.0) | 0.516 |
| Hypertension | 30(53.6) | 13(48.1) | 43(51.8) | 71(58.7) | 0.389 |
| Atrial Fibrillation | 6(10.7) | 5(18.5) | 11(13.3) | 22(18.2) | 0.440 |
| COPD | 13(23.2) | 6(22.2) | 19(22.9) | 30(24.8) | 0.868 |
| Prior PCI | 3(5.4) | 5(18.5) | 8(9.6) | 14(11.6) | 0.819 |
| Prior CABG | 0(0.0) | 0(0.0) | 0(0.0) | 1(0.8) | 1.000 |
| Prior MI | 0(0.0) | 1(3.7) | 1(1.2) | 1(0.8) | 1.000 |
| Prior Stroke | 0(0.0) | 1(3.7) | 1(1.2) | 10(8.3) | **0.030** |
| Syncope | 6(10.7) | 3(11.1) | 9(10.8) | 10(8.3) | 0.626 |
| Medication on admission |  |  |  |  | 0.592 |
| Anticoagulation | 5(8.9) | 3(11.1) | 8(9.6) | 14(11.6) |  |
| Antiplatelet | 28(50.0) | 11(40.7) | 39(47.0) | 63(52.1) |  |
| No antithrombosis | 23(41.1) | 13(48.1) | 36(43.4) | 44(36.4) |  |
| Pre TTE data |  |  |  |  |  |
| Mean Gradient (mmHg) | 59.7±23.0 | 55.8±17.1 | 58.4±21.2 | 56.3±37.2 | 0.307* |
| AVA (cm<sup>2</sup>) | 0.50±0.18 | 0.55±0.18 | 0.52±0.18 | 0.66±0.23 | **0.000\*** |
| Max velocity (m/s) | 5.01±0.93 | 4.87±0.70 | 4.96±0.86 | 4.75±0.72 | 0.065 |
| AR moderate/severe | 6(10.7) | 5(18.5) | 11(13.3) | 57(47.1) | **0.000** |
| EF | 53.6±14.5 | 55.9±11.6 | 54.4±13.6 | 55.2±14.1 | 0.547* |
| Pre CT data |  |  |  |  |  |
| Area (mm<sup>2</sup>) | 464.8±89.8 | 452.3±91.6 | 461.6±89.6 | 459.5±96.2 | 0.709* |
| Perimeter (mm) | 78.0±7.2 | 76.7±6.9 | 77.7±7.1 | 77.0±7.8 | 0.561* |
| STJ diameter (mm) | 31.80±3.68 | 28.74±4.11 | 31.01±3.99 | 28.47±5.29 | **0.005** |
| STJ height (mm) | 25.07±5.77 | 22.16±4.10 | 24.32±5.50 | 22.67±4.33 | 0.141* |
| Ascending aorta diameter at 4cm (mm) | 38.89±3.37 | 37.09±2.31 | 38.42±3.21 | 36.64±3.84 | **0.009** |
| Max ascending aorta diameter (mm) | 43.86±3.99 | 39.22±2.77 | 42.66±4.22 | 38.80±4.66 | **0.000** |
| RCA height (mm) | 18.1±5.7 | 15.6±2.6 | 17.4±5.2 | 16.6±4.0 | 0.694* |
| LM height (mm) | 17.7±4.6 | 15.5±2.2 | 17.2±4.2 | 14.3±3.0 | **0.000\*** |
| Aortic Root Angle | 52.8±9.8 | 52.7±8.4 | 52.8±9.4 | 51.1±8.8 | 0.338 |
| Calcification grade (II+) | 46(82.1) | 21(77.8) | 67(80.7) | 72(59.5) | **0.002** |

\* Mann-Whitney U test was used.
Data are presented as mean ± SD or no. (%). AS, aortic stenosis; AVA, aortic valve area; AR, aortic regurgitation; BAV, bicuspid aortic valve; BMI, body mass index; CABG, coronary artery bypass grafting; COPD, chronic obstructive pulmonary disease; EF, ejection fraction; GI, Gastrointestinal; LM, left main artery; MI, myocardial infarction; NYHA, New York Heart Association; PCI, percutaneous coronary intervention; PPI, permanent pacemaker implantation; PVD, peripheral vascular disease; RCA, right coronary artery; STJ, sino-tubular Junction; STS, Society of Thoracic Surgeons; TAV, tricuspid aortic valve.

The aortic valve area (AVA) (0.52 ± 0.18 cm^2^ vs. 0.66 ± 0.23 cm^2^, p = 0.000) and prevalence of moderate or severe aortic regurgitation (13.3% vs. 47.1%, p = 0.000) was lower in BAV patients. Sino-tubular junctions (STJ) were larger (31.01 ± 3.99 mm vs. 28.47 ± 5.29 mm, p = 0.005) in the BAV group compared to the TAV group. Larger ascending aorta diameters were observed in BAV patients not only at 40 mm from the annulus (38.42 ± 3.21 mm vs. 36.64 ± 3.84 mm, p = 0.009) but also at the max ascending aorta plane (42.66 ± 4.22 mm vs. 38.80 ± 4.66 mm, p = 0.000). BAV patients had higher left main coronary artery height (17.2 ± 4.2 mm vs. 14.3 ± 3.0 mm, p = 0.000) and more severe calcification (Calcification more than grade I, 80.7% vs. 59.5%, p = 0.002). There were no differences between the two groups in local anesthetic consideration, and pre-dilatation times during the TAVR procedure. The prevalence of post-dilatation was higher in the BAV group than that of the TAV group (63.9% vs. 48.8%, p = 0.045). Procedure duration (65.0 ± 46.2 min vs. 60.7 ± 33.8 min, p = 0.153) tended to be longer in BAV patients compared to TAV patients without significance. Moreover, a higher proportion of BAV patients adopted self-expandable devices, while more balloon-expandable were implanted in TAV patients (BAV vs. TAV: Edwards Sapien, 3.6% vs. 22.3%; Venus A, 75.9% vs. 61.2%; MicroPort, 0.0% vs. 1.7%; Lotus, 0.0% vs. 2.5%; Others, 20.5% vs. 12.4%; p = 0.000). Echocardiographic and computed tomographic baseline were listed in **Table 1**. Procedural data were listed in **Table 2**.

**Table 2.** Procedural data and DW-MRI findings following TAVR

|  | BAV |  |  | TAV | P value |
| --- | --- | --- | --- | --- | --- |
|  | BAV (Type 0)<br>n = 56 | BAV (Type 1)<br>n = 27 | Total<br>n = 83 | n = 121 |  |
| Procedural data |  |  |  |  |  |
| Local Anesthetic | 47(83.9) | 24(88.9) | 71(85.5) | 106(87.6) | 0.679 |
| Pre-dilatation |  |  |  |  | 0.121 |
| No pre-dilatation | 0(0.0) | 0(0.0) | 0(0.0) | 6(5.0) |  |
| Pre-dilatation once | 48(85.7) | 14(51.9) | 62(74.7) | 89(73.9) |  |
| Multiple pre-dilatation | 8(14.3) | 13(48.1) | 21(25.3) | 26(21.5) |  |
| Post-dilatation | 38(67.9) | 15(55.6) | 53(63.9) | 59(48.8) | **0.045** |
| Duration of procedure (min) | 69.7±32.8 | 55.4±26.4 | 65.0±46.2 | 60.7±33.8 | 0.153* |
| Contrast (ml) | 136.3±50.2 | 128.5±36.6 | 133.8±46.2 | 125.8±45.2 | 0.083* |
| Valve type |  |  |  |  | **0.000** |
| Edwards | 0(0.0) | 3(11.1) | 3(3.6) | 27(22.3) |  |
| Venus A | 45(80.4) | 18(66.7) | 63(75.9) | 74(61.2) |  |
| MicroPort | 0(0.0) | 0(0.0) | 0(0.0) | 2(1.7) |  |
| Lotus | 0(0.0) | 0(0.0) | 0(0.0) | 3(2.5) |  |
| Others | 11(19.6) | 6(22.2) | 17(20.5) | 15(12.4) |  |
| Patients with new lesions | 48(85.7) | 22(81.5) | 70(84.3) | 92(76.0) | 0.163 |
| Lesion location(patients) |  |  |  |  | 0.203 |
| Left hemisphere | 4(8.3) | 4(18.2) | 8(11.4) | 20(21.7) |  |
| Right hemisphere | 9(18.8) | 3(13.6) | 12(17.1) | 17(18.5) |  |
| Bilateral lesions | 35(72.9) | 15(68.2) | 50(71.4) | 55(59.8) |  |
| Lesion size(patients) |  |  |  |  | **0.005** |
| ≤1 cm<sup>3</sup> | 35(72.9) | 15(68.2) | 50(71.4) | 82(89.1) |  |
| >1 cm<sup>3</sup> | 13(27.1) | 7(31.8) | 20(28.6) | 10(10.9) |  |
| Total number of lesions | 333 | 139 | 472 | 423 |  |
| Number of lesions | 5.95±6.32 | 5.15±6.09 | 5.69±6.22 | 3.50±4.16 | **0.008\*** |
| Number of lesions<sup>#</sup> | 4.0(1.0-8.0) | 3.0(1.0-8.0) | 4.0(1.0-8.0) | 2.0(1.0-5.0) |  |
| Lesion location(number) |  |  |  |  |  |
| ACA lesions | 0.77±1.36 | 1.00±1.69 | 0.84±1.47 | 0.55±1.21 | 0.107* |
| ACA/MCA | 1.11±1.67 | 1.00±1.73 | 1.07±1.68 | 0.50±1.05 | **0.007\*** |
| MCA lesions | 1.50±2.22 | 1.37±1.74 | 1.46±2.07 | 0.98±1.84 | **0.039\*** |
| MCA/PCA | 0.16±0.66 | 0.11±0.42 | 0.14±0.59 | 0.08±0.33 | 0.808* |
| PCA lesions | 1.25±2.33 | 0.96±1.77 | 1.16±2.16 | 0.61±1.16 | 0.137* |
| VA/BA lesions | 1.16±1.35 | 0.70±1.33 | 1.01±1.35 | 0.77±1.44 | **0.033\*** |
| Total lesion volume(mm<sup>3</sup>) | 1098±2648 | 691±1130 | 966±2268 | 1159±9144 | **0.008\*** |
| Total lesion volume(mm<sup>3</sup>)<sup>#</sup> | 335(70-898) | 250(70-1070) | 290(70-930) | 140(35-480) |  |
| Volume/number(mm<sup>3</sup>) | 147.4±377.9 | 93.8±92.2 | 129.9±314.9 | 362.8±3053.4 | **0.037\*** |
| Volume/number(mm<sup>3</sup>)<sup>#</sup> | 68.0(41.1-114.3) | 80.0(55.0-125.0) | 70.0(45.0-115.0) | 57.5(24.5-93.0) |  |
| Time of post-procedural DW-MRI (days) | 5.7±2.5 | 6.3±3.8 | 5.9±2.9 | 5.6±2.7 | 0.750* |

\* Mann-Whitney U test was used.
<sup>#</sup> Data was presented by median(interquartile range).
Data are presented as mean ± SD or no. (%). ACA, anterior cerebral artery; BA, basilar artery; BAV, bicuspid aortic valve; DW-MRI, diffusion-weighted magnetic resonance imaging; MCA, middle cerebral artery; PCA, posterior cerebral artery; TAV, tricuspid aortic valve; VA, vertebral artery.

There were 2 BAV patients experienced a non-disabling stroke while 2 patients in the TAV group had a disabling stroke during their hospital stays. No significant difference was found in other peri-procedural complications. 1 death occurred in the TAV group in hospital. Peri-procedural and follow-up clinical outcomes were listed in **Supplementary Table 1**.

### DW-MRI Data

DW-MRI was performed several days after TAVR in both BAV group and TAV group. There was no difference between the two groups in the number of days after TAVR the DW-MRI was performed. The incidence of new ischemic cerebral lesions in BAV patients after TAVR was not different from those in TAV patients (84.3% vs. 76.0%, p = 0.163). However, the number of lesions was higher in the BAV group (5.69 ± 6.22 vs. 3.50 ± 4.16, p = 0.008) and the proportion of patients with lesion size greater than 1cm^3^ was higher in BAV patients (28.6% vs. 10.9%, p = 0.005) (**Table 2**). Moreover, the number of new lesions was higher in BAV patients in the ACA/MCA zone (1.07 ± 1.68 vs. 0.50 ± 1.05, p = 0.007), MCA (1.46 ± 2.07 vs. 0.98 ± 1.84, p = 0.039), and in the VA/BA lesions zone (1.01 ± 1.35 vs. 0.77 ± 1.44, p = 0.033) (**Table 2**). The total volume of new lesions [290(70-930) mm^3^ vs. 140(35-480) mm^3^, p = 0.008] and volume per lesion [70.0(45.0-115.0) mm^3^ vs. 57.5(24.5-93.0) mm^3^, p = 0.037] were significantly bigger in the BAV group when compared with the TAV group (**Central Illustration**). The distribution of the number of lesions in ACA, ACA/MCA, MCA, MCA/PCA, PCA, and VA/BA zones were comparable in BAV and TAV patients without statistical significance (**Figure 2, Supplementary Table 2**). Age (p = 0.010), Procedure duration (p = 0.000) and BAV (p = 0.008) were independent predictors of new ischemic lesion occurrence in the multivariable linear regression (**Table 3**).

**Figure 2.**
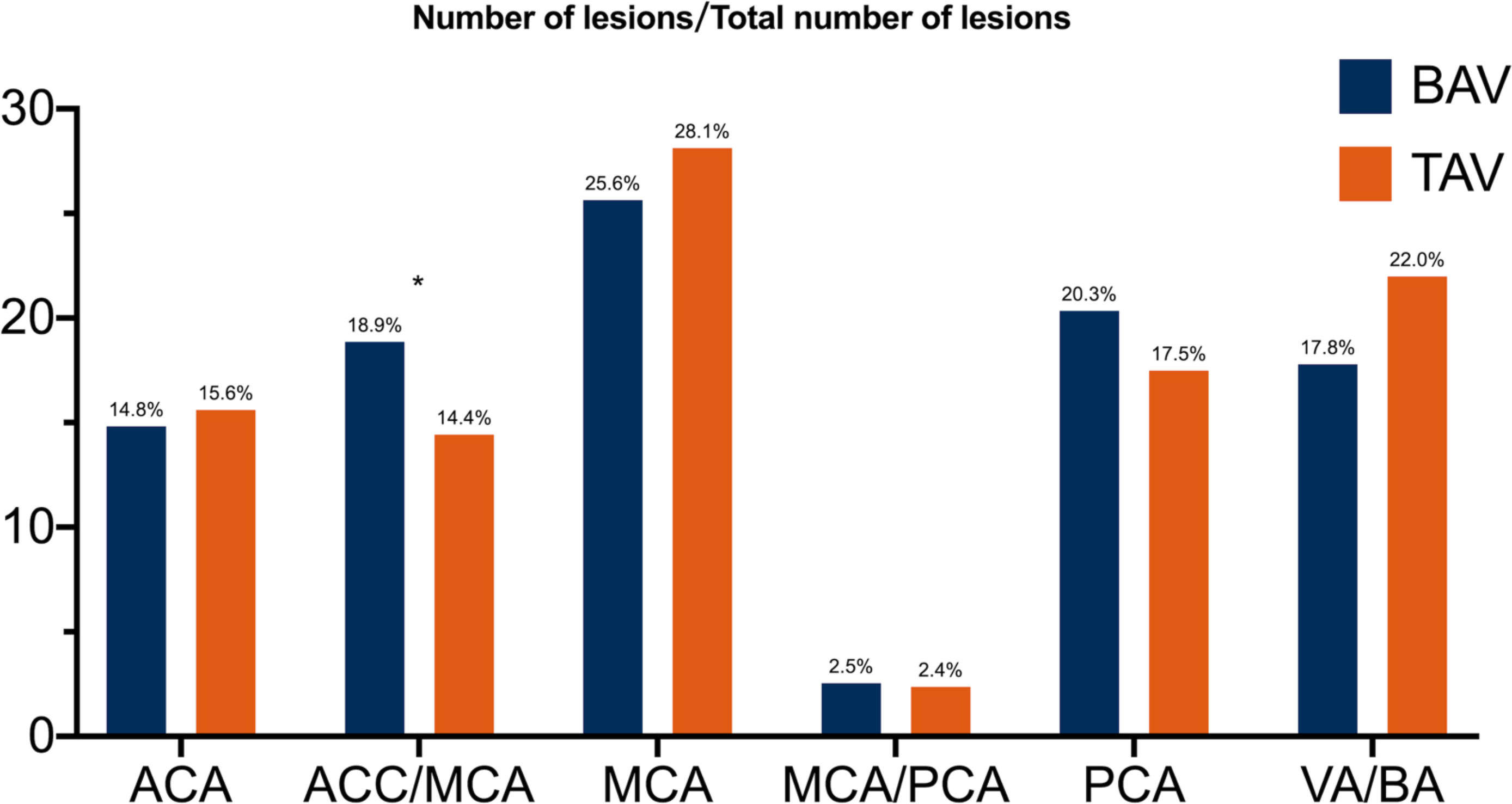
The distribution of the new cerebral ischemic lesions in bicuspid and tricuspid aortic valve patients

**Table 3.** Predictor of number of lesions

| Predictors | Univariate linear regression |  |  | Multivariate linear regression* |  |  |
| --- | --- | --- | --- | --- | --- | --- |
|  | Coefficients | Standard error | P value | Coefficients | Standard error | P value |
| Age (yrs) | 0.095 | 0.053 | 0.075 | 0.149 | 0.057 | 0.010 |
| BMI (kg/m<sup>2</sup>) | -0.116 | 0.111 | 0.299 |  |  |  |
| STS | -0.004 | 0.092 | 0.965 |  |  |  |
| Prior Stroke | -0.794 | 1.615 | 0.624 |  |  |  |
| AVA | -3.807 | 1.648 | 0.022 |  |  |  |
| Max velocity (m/s) | 0.666 | 0.463 | 0.152 |  |  |  |
| Multiple Pre-dilatation | 0.494 | 0.778 | 0.526 |  |  |  |
| Post-dilatation | 1.319 | 0.728 | 0.071 |  |  |  |
| Duration of procedure | 0.057 | 0.011 | 0.000 | 0.059 | 0.011 | 0.000 |
| Contrast | 0.027 | 0.009 | 0.002 |  |  |  |
| Calcification Grade | 1.110 | 0.779 | 0.156 |  |  |  |
| LM height | -0.007 | 0.128 | 0.954 |  |  |  |
| STJ diameter | 0.051 | 0.102 | 0.615 |  |  |  |
| Ascending aorta diameter (4cm) | 0.068 | 0.136 | 0.618 |  |  |  |
| Ascending aorta diameter (Max) | 0.015 | 0.102 | 0.888 |  |  |  |
| Self vs. Balloon expandable Valve | -0.369 | 0.991 | 0.710 |  |  |  |
| BAV vs. TAV | -2.191 | 0.727 | 0.003 | -2.108 | 0.784 | 0.008 |

\*Stepwise selection was used to do the multivariable linear regression
AS, aortic stenosis; AVA, aortic valve area; BAV, bicuspid aortic valve; LM, left main artery; STJ, sino-tubular Junction; STS, Society of Thoracic Surgeons; TAV, tricuspid aortic valve.

## Discussion

The main findings of the present study are that BAV patients may experience a higher number of cerebral ischemic lesions. Moreover, the BAV patients are also associated with larger size of new lesions after TAVR, especially in the ACA/MCA, MCA, and VB/BA lesions zone, accompanied with frequent deployment of self-expandable devices, longer procedure duration, and more frequent post-dilatation.

The bicuspid aortic valve is often heavily calcified and accompanied by raphe (fusion between adjacent cups) and concomitant aortopathy (dilatation of the ascending aorta), which may require additional surgical treatment of the aorta. As TAVR in BAV patients presents both anatomical and clinical challenges, BAV patients were usually excluded from many major randomized TAVR clinical trials (1–6). Until now, only a few studies have reported the safety and efficacy of TAVR in BAV patients(11–14). This study found that the stroke rate was 2.4% in BAV patients and 1.7% in TAV patients. In consistent with the present findings, Makkar RR et al. also reported that younger and intermediate-to-low risk bicuspid AS patients undergoing TAVR had higher prevalence of stroke when compared to tricuspid AS patients, which was similar with our results (10).

The high reported incidence of new cerebral ischemic lesions on post-TAVR DW-MRI had raised the concern of the physicians in the BAV patients whose multiple balloon valvuloplasty perhaps would induce more native valve debris (15–17, 19, 20). In the present study, we had indeed found the higher number and bigger of new cerebral ischemic lesions in BAV patients than TAV patients, even though the stroke rates were not significantly different between the two groups.

### Procedural Mechanism of Cerebral Ischemic Lesions

In the present study, TAVR in BAV patients was feasible, but also led to a longer procedure duration which may be related to higher post-dilation frequency in BAV group. A previous study identified balloon valvuloplasty and actual valve positioning and deployment as the primary cause of cerebral embolization during TAVR procedure by transcranial doppler (25). Histopathological analysis of debris captured by a dual filter–based embolic protection device (Montage Dual Filter System, Claret Medical, Inc., Santa Rosa, California) in 81 TAVR patients revealed that these tissues originated from the native aortic valve leaflets, aortic wall, or left ventricular myocardium and were more frequent with the use of balloon-expandable systems and more oversizing (26). It is possible that essential manipulations like crossing a calcified aortic valve and subsequent instrumentation within the aortic root including valve positioning and placement are equally responsible for dislodgment of material from the aortic valve and aorta (26). Therefore, it is reasonable that the procedural complexity in BAV patients with longer procedure duration and higher post-dilation frequency may add to the risk of cerebral injury by increasing the number of new ischemic lesions after TAVR in DW-MRI. However, we didn’t find the obvious evidence between the pre-dilatation times during the TAVR procedure and new ischemic lesions in DW-MRI after TAVR.

### Predictive Factors of Cerebral Ischemic Lesions

In the current study, we systemically analyzed the new ischemic lesions, and found that BAV patients had a higher proportion of larger lesions and a greater number of lesions, especially in the ACA/MCA, MCA and VA/BA lesions zone. Patients with BAV are usually younger with larger lesion size as examined by DW-MRI. In contrast, older patients (TAV population) usually have a higher likelihood of cerebral atrophy, which may result in smaller size lesions. Our data also suggest that BAV patients had more prevalence of lesions larger than 1 cm^3^, but the total lesion volume presented by mean ± standard error was lower. This may explain the presence of the outlier caused by disabling stroke in TAV patients, because disabling strokes can increase the value of the total volume but have little effect on the proportion of large-scale lesions. Therefore, the total lesion volume and the ratio of volume and number was lower in TAV patients when data were presented by median (interquartile range).

The previous studies showed that age, STS score, duration of procedure, fluoroscopy time, and valve type were related to the ischemic lesions in DW-MRI (24). In our univariable linear regression analysis, we identified that Aortic Valve Area (AVA), duration of procedure, Contrast, and BAV were predictors of the number of new lesions. The multivariable linear regression analysis revealed that age, procedure duration, and BAV were independent factors influencing the number of new lesions. Procedural complexity due to complex BAV anatomy may necessitate post-dilatation in BAV patients. The higher proportion of post-dilatation, valve manipulation, and rapid pacing may increase the duration of procedure and the risk of hypoperfusion, which may also increase the number of lesions. Moreover, heavy calcification could also increase the risk of small debris originating from the calcified native aortic valve, causing heavier damage in the younger BAV patients.

### Cerebral Ischemic Lesions and Protection Devices in TAVR

New cerebral ischemic lesions were found in 74%-100% of patients after TAVR in DW-MRI^15–17^. Though some studies show that new ischemic lesions are not linked to apparent neurological symptoms, there is evidence that perioperative ischemia may increase the risk of cognitive function and long-term dementia (27, 28).

In the randomized CLEAN-TAVI (CLaret Embolic Protection ANd TAVI Trial) study, the number of DW-MRI cerebral ischemic lesions decreased after use of the cerebral protection devices in patients undergoing TAVR and significantly improved short-term neurological outcome (29). Moreover, the DEFLCT trial also demonstrated that TriGuard Embolic Deflection Device during TAVR procedure could reduce ischemic brain volume, and subjects suffered fewer neurologic deficits (30).

### Clinical Implications

The present study is the first to provide insights into the risk of brain injury in DW-MRI for the TAVR procedure in BAV patients. Further studies are necessary to show whether TAVR is suitable for BAV patients when considering brain injury complications. Cerebral embolic protection devices may be recommended for TAVR, especially in BAV patients, to avoid cerebral ischemic lesions that potentially deteriorate neurological and cognitive function.

### Study Limitations

One limitation of the present study is that this is a single-center, non-randomized study, and therefore needs to be confirmed by a larger randomized clinical trial. Even though the prevalence of patients with prior atrial fibrillation or new onset atrial fibrillation were similar between two groups, cardiac thrombus associated with atrial fibrillation should not be excluded as a potential contributor to micro thrombosis. Moreover, new generation devices could be used for TAVR in the clinical trial to see whether this result could be replicated. The present study still requires long-term follow-up to complete the mortality, stroke, neurological, and cognitive function assessment.

## Conclusions

A significantly higher frequency of larger new cerebral ischemic lesions after TAVR are found in BAV patients and are mostly located in the ACA/MCA, MCA and VA/BA lesions zone, which needs to be further confirmed by future studies.

## Data Availability

All data referred to in the manuscript will be available after the manuscript published.

## PERSPECTIVES

**WHAT IS KNOWN?** Silent cerebral ischemic lesions are present after TAVR in the majority of patients. Given the higher prevalence of bicuspid aortic valve stenosis in younger patients, coupled with the TAVR indication shift toward younger and lower risk patients, more and more studies have evaluated and preliminary validated the safety and efficacy of TAVR in BAV patients.

**WHAT IS NEW?** TAVR in BAV patients may encounter more severe brain injuries not only due to greater number of lesions but also due to larger lesion size. Therefore, TAVR in BAV patients should be more careful and cautious to avoid cerebral ischemic lesions that potentially deteriorate neurological and cognitive function.

**WHAT IS NEXT?** This finding has to be proven in a randomized trial. Moreover, cerebral embolic protection devices may be recommended for TAVR in BAV patients.

**Abbreviations**
AS: aortic stenosis
BAV: bicuspid aortic valve
CT: computed tomography
DW-MRI: diffusion-weighted magnetic resonance imaging
HU: Hounsfield unit
TAV: tricuspid aortic valve
TAVR: transcatheter aortic valve replacement
TTE: transthoracic echocardiography

---

Central Illustration. The brain injury in bicuspid and tricuspid aortic valve patients

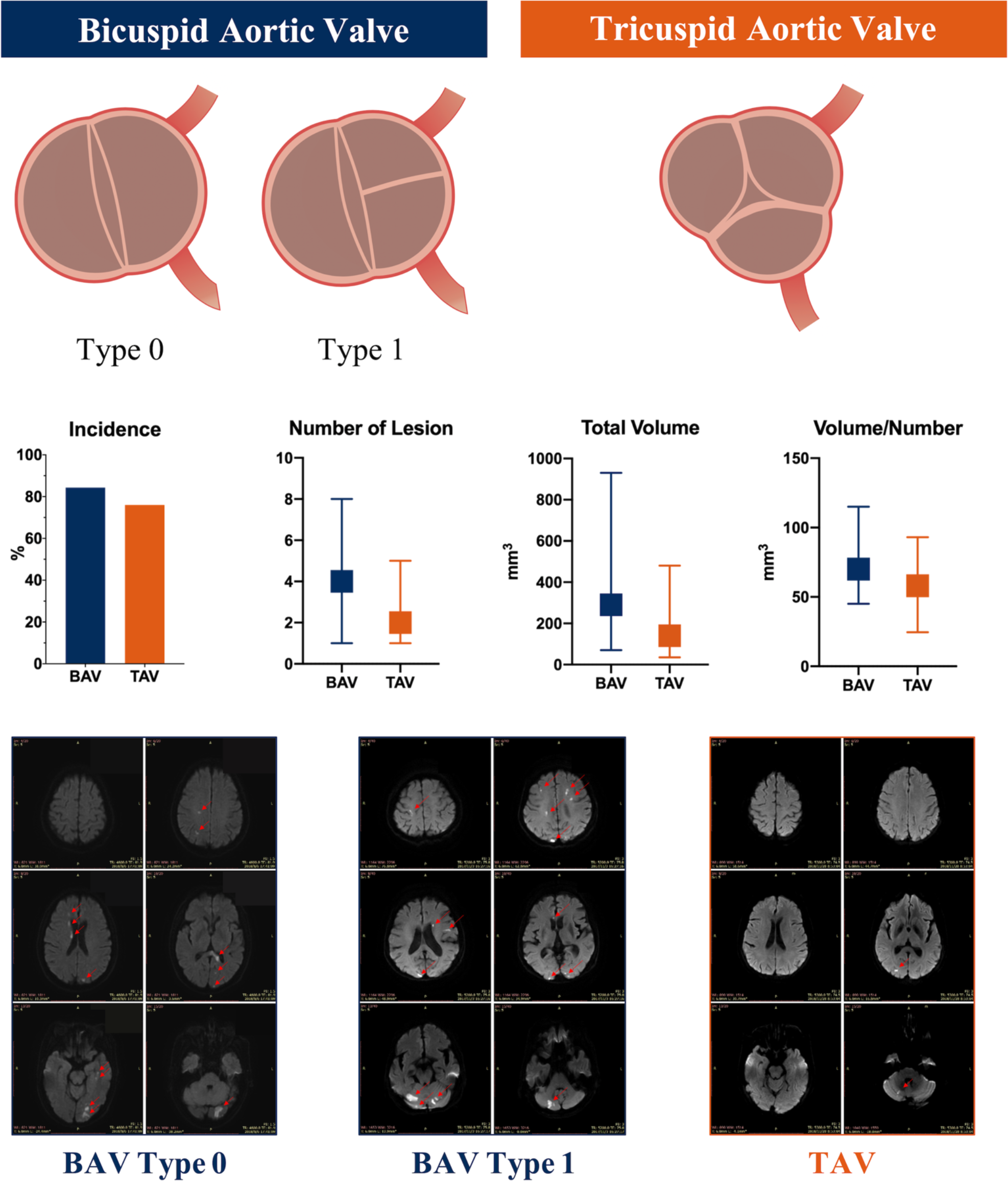

**Supplementary Table 1.** Peri-procedural and follow-up clinical outcomes

|  | BAV |  |  | TAV | P value |
| --- | --- | --- | --- | --- | --- |
|  | BAV (Type 0)<br>n = 56 | BAV (Type 1)<br>n = 27 | Total<br>n = 83 | n = 121 |  |
| Mortality |  |  |  |  |  |
| In-hospital | 0(0.0) | 0(0.0) | 0(0.0) | 1(0.8) | 1.000 |
| 30-day | 0(0.0) | 0(0.0) | 0(0.0) | 1(0.8) | 1.000 |
| Stroke | 2(3.6) | 0(0.0) | 2(2.4) | 2(1.7) | 0.704 |
| Disabling stroke | 0(0.0) | 0(0.0) | 0(0.0) | 2(1.7) | 0.515 |
| Non-disabling stroke | 2(3.6) | 0(0.0) | 2(2.4) | 0(0.0) | 0.164 |
| Vascular Complication | 5(8.9) | 2(7.4) | 7(8.4) | 10(8.3) | 1.000 |
| New Af | 1(1.8) | 0(0.0) | 1(1.2) | 4(3.3) | 0.412 |
| New Pacemaker | 0(0.0) | 0(0.0) | 0(0.0) | 4(3.3) | 0.147 |
| Medication |  |  |  |  | 0.953 |
| Anticoagulation | 8(14.3) | 5(18.5) | 13(15.7) | 21(17.4) |  |
| Antiplatelet | 46(82.1) | 22(81.5) | 68(81.9) | 96(79.3) |  |
| No antithrombosis | 2(3.6) | 0(0.0) | 2(2.4) | 4(3.3) |  |

**Supplementary Table 2.** Detail distribution of the new ischemic lesions

|  | BAV<br>n = 83 | TAV<br>n = 121 | P |
| --- | --- | --- | --- |
| Total number of lesions | 472 | 423 |  |
| Lesion location, number* |  |  | 0.290 |
| ACA lesions | 70(14.8)<br>[-0.3] | 66(15.6)<br>[0.3] |  |
| ACC/MCA | 89(18.9)<br>[1.8] | 61(14.4)<br>[-1.8] |  |
| MCA lesions | 121(25.6)<br>[-0.8] | 119(28.1)<br>[0.8] |  |
| MCA/PCA | 12(2.5)<br>[0.2] | 10(2.4)<br>[-0.2] |  |
| PCA lesions | 96(20.3)<br>[1.1] | 74(17.5)<br>[-1.1] |  |
| VA/BA lesions | 84(17.8)<br>[-1.6] | 93(22.0)<br>[1.6] |  |

\*Adjusted residuals appear in square brackets below observed frequencies and when it's difference > 3.0 or < -3.0 means significant
ACA, anterior cerebral artery; BA, basilar artery; BAV, bicuspid aortic valve; MCA, middle cerebral artery; PCA, posterior cerebral artery; TAV, tricuspid aortic valve; VA, vertebral artery.

